# Resilience and social support improve mental health and quality of life in patients with post-COVID-19 syndrome

**DOI:** 10.1101/2023.02.07.23285620

**Authors:** Petros Galanis, Aglaia Katsiroumpa, Irene Vraka, Katerina Kosiara, Olga Siskou, Olympia Konstantakopoulou, Theodoros Katsoulas, Parisis Gallos, Daphne Kaitelidou

## Abstract

The effects of post-COVID-19 syndrome on patients’ life are significant. As there is no prior study available, we investigated the impact of resilience and social support on anxiety, depression, and quality of life among patients with post-COVID-19 syndrome. We conducted a cross-sectional study with a convenience sample. The measures included demographic and clinical characteristics of patients, the Brief Resilience Scale, the Multidimensional Scale of Perceived Social Support, the Patient Health Questionnaire-4, and the EuroQol-5D-3L. Multivariable analysis identified that resilience and social support reduced anxiety and depression among our patients. Also, we found a significant positive relationship between resilience and social support, and quality of life. In conclusion, our findings suggest that resilience and social support can be protective by reducing anxiety and depression, and improving quality of life among patients with post-COVID-19 syndrome. Policy makers should develop and implement healthcare management programs to provide psychological support to these patients.

## Introduction

Post-COVID-19 syndrome, also known as long COVID-19, is defined by the World Health Organization (WHO) as the condition where symptoms are present three months after SARS-CoV-2 infection and cannot be explained by alternative diagnoses (World Health Organization, 2023a). Fatigue, shortness of breath, sleep disorders, and cognitive dysfunction are the most common symptoms in patients with post-COVID-19 syndrome, while over 200 long-term different symptoms are identified in the literature (Alkodaymi et al., 2022; Nasserie et al., 2021). A large proportion of COVID-19 patients has also post-COVID-19 syndrome. For example, according to a prediction model from WHO, more than 17 million people across the WHO European Region may have experienced post-COVID-19 syndrome during first two years of the pandemic (World Health Organization, 2023b). Moreover, a meta-analysis with a total of 1.2 million symptomatic COVID-19 cases found that the fatigue cluster of long COVID-19 occurred in 51.0% of cases, the respiratory cluster occurred in 60.4% of cases, and the cognitive cluster occurred in 35.4% of cases (Global Burden of Disease Long COVID Collaborators et al., 2022). Additionally, at >12-month follow-up, 41%, 31%, 30%, 22%, and 22% of confirmed SARS-CoV-2 cases continued to experience fatigue, dyspnea, sleep disorder, myalgia, and cognitive impairment respectively (Alkodaymi et al., 2022; Ceban et al., 2022).

Impact of post-COVID-19 syndrome on mental health of patients is large. The European Centre for Disease Prevention and Control (ECDC) performed a meta-analysis and found that prevalence of anxiety, depression, and post-traumatic stress disorder in patients with post-COVID-19 syndrome recruited in the community settings was 17.2%, 17.3%, and 20.6% respectively (European Centre for Disease Prevention and Control, 2022). Prevalence of mental health issues was even higher for patients recruited in the hospital settings, i.e., prevalence of anxiety was 27.5%, and depression was 23.3%. A similar systematic review found that anxiety (range from 6.5% to 63% across the studies), depression (range from 4% to 31%), and post-traumatic stress disorder (range from 12.1% to 46.9%) were the most common mental health issues in patients with post-COVID-19 syndrome (Shanbehzadeh et al., 2021).

Moreover, post-COVID-19 syndrome affects patients’ quality of life since a meta-analysis found that the prevalence of poor quality of life was 59% (Malik et al., 2022). Additionally, among patients with post-COVID-19 syndrome, 41.5% reported extreme pain/discomfort, 37.5% reported extreme anxiety/depression, 36% reported extreme problems on mobility, 28% reported extreme problems on usual activities, and 8% reported extreme problems on self-care (Malik et al., 2022).

The positive impact of resilience and social support during the pandemic has already proven. In particular, a meta-analysis found a significant negative relationship between resilience and psychological distress among COVID-19 patients (Jeamjitvibool et al., 2022). Moreover, a review including workers found that resilience was associated with lower levels of anxiety, depression and burnout (Finstad et al., 2021). Similarly, a systematic review found that resilience and social support among healthcare workers improved mental and psychological health outcomes (Labrague, 2021). Additionally, low perceived social support was a risk factor of adverse psychological states of COVID-19 patients, such as stress, anxiety, and depression (Dong et al., 2021). Also, social support seemed very important during the first wave of pandemic due its negative correlation with psychological pressure, mental discomfort and anxiety (Filindassi et al., 2022).

Until now, research has only explored the relationship between a few demographic and clinical characteristics of patients with post-COVID-19 syndrome and mental health and quality of life. Greater anxiety and depression were reported in female patients, patients with psychiatric history, and patients with intensive care unit admission (Renaud-Charest et al., 2021; Shanbehzadeh et al., 2021). Moreover, quality of life was lower among patients admitted to intensive care, and patients with higher levels of fatigue (Malik et al., 2022). However, the available studies have not investigated the influence of psychosocial factors on patients’ life. Thus, the aim of our study was to explore the impact of resilience and social support on anxiety, depression, and quality of life among patients with post-COVID-19 syndrome.

## Materials and Methods

### Study design and population

We conducted an online cross-sectional study in Greece between November 2022 and January 2023. First, we created an online version of the study questionnaire using Google forms. Then, we posted the questionnaire on the Facebook page of the Long COVID Greece patients’ society after administrators’ approval (*Long COVID Greece*, 2023). Patients can participate in our study by clicking on the link of the questionnaire. Therefore, we recruited patients with post-COVID-19 syndrome via convenience sampling. Response rate was unknown since we cannot measure the number of patients that clicked on the link of the study questionnaire but did not complete it. The Long COVID Greece patients’ society is a member of a European network of Long COVID patient associations, i.e., Long COVID Europe (*Long COVID Europe*, 2023). The Long COVID Greece patients’ society is a non-profit organization and is created by patients with post-COVID-19 syndrome.

Participation in our study was allowed if the following criteria were met: adults aged ≥18 years; individuals who understand the Greek language since the study questionnaire was in Greek; previous confirmed SARS-CoV-2 infection with molecular test; patients who presented symptoms and signs consistent with COVID-19 for ≥12 weeks after the COVID-19 diagnosis and were not attributable to alternative diagnoses in order to meet the definition of post-COVID-19 syndrome (Shah et al., 2021; World Health Organization, 2023a).

Minimum sample size required 121 patients considering a low effect size (f^2^=0.11), the power as 95%, the alpha level as 5%, and the number of independent variables as 10 (Lorah, 2018).

### Demographic and clinical characteristics

We measured gender (females or males), age (continuous variable), hospitalization in COVID-19 ward (no or yes), hospitalization in COVID-19 intensive care unit (no or yes), duration of COVID-19 symptoms (continuous variable), anxiety disorders before post-COVID-19 syndrome (no or yes), depression before post-COVID-19 syndrome (no or yes).

### Resilience

We measured patients’ resilience with the Brief Resilience Scale (BRS) (Smith et al., 2008). BRS consists of six items, e.g. “I tend to bounce back quickly after hard times”. Answers are on a five-point Likert scale, i.e., 1: strongly disagree, 2: disagree, 3: neutral, 4: agree, 5: strongly agree. Total score is calculated dividing the total sum by the total number of questions answered. Thus, a total score from 1 to 5 is produced with higher values indicating higher levels of resilience.

BRS is translated and validated in Greek language (Kyriazos et al., 2018). In our study, Cronbach’s coefficient alpha for the BRS was 0.878.

### Social support

We used the Multidimensional Scale of Perceived Social Support (MSPSS) to measure family support, friends support, and significant others support (Zimet et al., 1988). MSPSS consists of 12 items and answers are on a seven-point Likert scale from strongly disagree to strongly agree. Each factor includes four items and takes a total score from 1 to 7. Improved scores on MSPSS indicate higher level of support. We used a reliable and valid Greek version of MSPSS in our study (Theofilou, 2015). We found that Cronbach’s coefficient alpha for the factor “family support” was 0.968, the factor “friends support” was 0.971, and the factor “significant others support” was 0.919.

### Mental health outcomes

We used the Patient Health Questionnaire-4 (PHQ-4) to measure anxiety and depression among patients with post-COVID-19 syndrome (Kroenke et al., 2009). PHQ-4 consists of four items: two items measure anxiety and two items measure depression. Answers on items take values from 0 (not at all) to 3 (almost every day). Thus, total score on anxiety and depression scales ranges from 0 to 6. Also, a total score on PHQ-4 is calculated with values from 0 to 12. Higher values indicate higher levels of anxiety and depression. Individuals with a total score from 0 to 2 are rated as normal, those with a total score from 3 to 5 are considered as experiencing mild distress, those with a total score from 6 to 8 are considered as experiencing moderate distress, and those with a total score from 9 to 12 are considered as experiencing severe distress. Moreover, total anxiety or depression score ≥3 is indicative of possible major anxiety or depression disorder.

We used the reliable and valid Greek version of the PHQ-4 (Karekla et al., 2012). In our study, Cronbach’s coefficient alpha for the PHQ-4 was 0.864, the anxiety scale was 0.854, and the depression scale was 0.849.

### Quality of life

We used the 3-level version of EQ-5D (EQ-5D-3L) to measure patients’ quality of life (Rabin & de Charro, 2001). The EQ-5D-3L includes the EQ-5D descriptive system and the EQ visual analogue scale (EQ VAS). The EQ-5D descriptive system includes five dimensions (i.e., mobility, self-care, usual activities, pain/discomfort and anxiety/depression) and each of them has three levels (i.e., no problems, some problems, and extreme problems). Answers on the five dimensions result into a 5-digit number that describes individual’s health state. This 5-digit number is then converted into a single summary index value using a set of weights which are produced from representative samples of people from general population. We used the set of weights which were produced from a study in Greece (Yfantopoulos, 1999). Higher EQ-5D-3L index values indicate higher level of quality of life. Moreover, individuals can self-rate their health status on the EQ VAS which is a vertical visual analogue scale with values from 0 (worst imaginable health state) to 100 (best imaginable health state). In our study, Cronbach’s coefficient alpha for the EQ-5D-3L was 0.791.

### Ethical issues

Patients with post-COVID-19 syndrome can participate in the study in an anonymous and voluntary basis. Moreover, we did not collect personal information data, e.g. name, insurance number, IP address, email address, etc. We informed patients about the study aim and design in the first page of the online questionnaire. Then, patients who gave their informed consent can access the full version of the questionnaire.

Our study protocol was approved by the Ethics Committee of Faculty of Nursing, National and Kapodistrian University of Athens (reference number; 420, 10 October 2022). Additionally, we applied the guidelines of the Declaration of Helsinki.

### Statistical analysis

We use numbers and percentages to present categorical variables. Moreover, we use mean, standard deviation, median, minimum value, and maximum value to present continuous variables. Resilience, family support, friends support, and significant others support were the independent variables, while anxiety, depression, and quality of life were the dependent variables. We considered demographic and clinical characteristics of patients with post-COVID-19 syndrome as possible confounders.

We used the Kolmogorov-Smirnov test and Q-Q plots to check the normality of the continuous variables. Because the dependent variables followed the normal distribution we applied the linear regression analysis in order to find out the impact of predictors. First, we performed a univariate linear regression analysis and then we constructed a multivariable linear regression model in order to eliminate confounding. In that case, we present unadjusted and adjusted coefficients beta, 95% confidence intervals (CI), and p-values. We did not include hospitalization in COVID-19 intensive care unit in the linear regression analysis due to the limited variability, i.e., only four patients have been hospitalized in COVID-19 intensive care unit. Independent variables with a p-value <0.05 in the final multivariable model were considered as statistically significant. We used the IBM SPSS 21.0 (IBM Corp. Released 2012. IBM SPSS Statistics for Windows, Version 21.0. Armonk, NY: IBM Corp.) for the analysis.

## Results

Study population included 122 patients with post-COVID-19 syndrome with a mean age of 44.8 years. Most of them were females (73%). Among patients, 17.2% have been hospitalized in COVID-19 ward and 3.3% in COVID-19 intensive care unit. Mean duration of COVID-19 symptoms was 11.6 months. Among patients, 18.9% reported that they have been diagnosed with an anxiety disorder and 11.5% with depression before post-COVID-19 syndrome. Demographic and clinical characteristics of patients with post-COVID-19 syndrome are presented in Table 1.

**Table 1.** Demographic and clinical characteristics of patients with post-COVID-19 syndrome (N=122).

| **Variables** | **N** | **%** |
| --- | --- | --- |
| Gender |  |  |
| Females | 89 | 73.0 |
| Males | 33 | 27.0 |
| Age (years), mean, standard deviation | 44.8 | 11.5 |
| Hospitalization in COVID-19 ward |  |  |
| No | 101 | 82.8 |
| Yes | 21 | 17.2 |
| Hospitalization in COVID-19 intensive care unit |  |  |
| No | 118 | 96.7 |
| Yes | 4 | 3.3 |
| Duration of COVID-19 symptoms (months), mean, standard deviation | 11.6 | 8.6 |
| Anxiety disorders before post-COVID-19 syndrome |  |  |
| No | 99 | 81.1 |
| Yes | 23 | 18.9 |
| Depression before post-COVID-19 syndrome |  |  |
| No | 108 | 88.5 |
| Yes | 14 | 11.5 |

Descriptive statistics for the scales in our study are shown in Table 2. Total PHQ score suggested that 32.8% (n=40) of patients with post-COVID-19 syndrome experienced severe psychological distress, 32.8% (n=40) experienced moderate distress, 23% (n=28) experienced mild distress, and 11.5% (n=14) had no distress. Moreover, 60.7% (n=74) of patients had anxiety score ≥3 and 69.7% (n=85) had depression score ≥3, indicating possible major anxiety or depression disorder.

**Table 2.** Descriptive statistics for the scales in our study.

| **Scale** | **Mean** | **Standard**<br>**deviation** | **Median** | **Minimum**<br>**value** | **Maximum**<br>**value** |
| --- | --- | --- | --- | --- | --- |
| Patient Health Questionnaire |  |  |  |  |  |
| Anxiety score | 3.22 | 1.78 | 3 | 0 | 6 |
| Depression score | 3.56 | 1.69 | 4 | 0 | 6 |
| EQ-5D-3L index value | 0.36 | 0.36 | 0.31 | -0.33 | 1 |
| EQ-5D-3L VAS | 54.10 | 21.71 | 55 | 0 | 100 |
| Brief Resilience Scale | 3.32 | 0.87 | 3.33 | 1.17 | 5 |
| Multidimensional Scale of Perceived Social Support |  |  |  |  |  |
| Family support | 5.41 | 1.61 | 5.75 | 1 | 7 |
| Friends support | 5.22 | 1.56 | 5.63 | 1 | 7 |
| Significant others support | 5.73 | 1.38 | 6 | 1 | 7 |

Mean EQ-5D-3L index value was 0.36 and mean EQ-5D-3L VAS was 54.1. Mean resilience score was 3.32 indicating a moderate level of resilience. Patients received more support from significant others (mean=5.73) than family (mean=5.41) and friends (mean=5.22).

Multivariable linear regression analysis identified that increased resilience (adjusted beta = -0.56, 95% CI = -0.94 to -0.17) and significant others support (adjusted beta = - 0.34, 95% CI = -0.64 to -0.03) reduced anxiety (Table 3). Also, we found that resilience (adjusted beta = -0.42, 95% CI = -0.80 to -0.05) and significant others support (adjusted beta = -0.39, 95% CI = -0.69 to -0.09) reduced depression in patients with post-COVID-19 syndrome (Table 4).

**Table 3.**
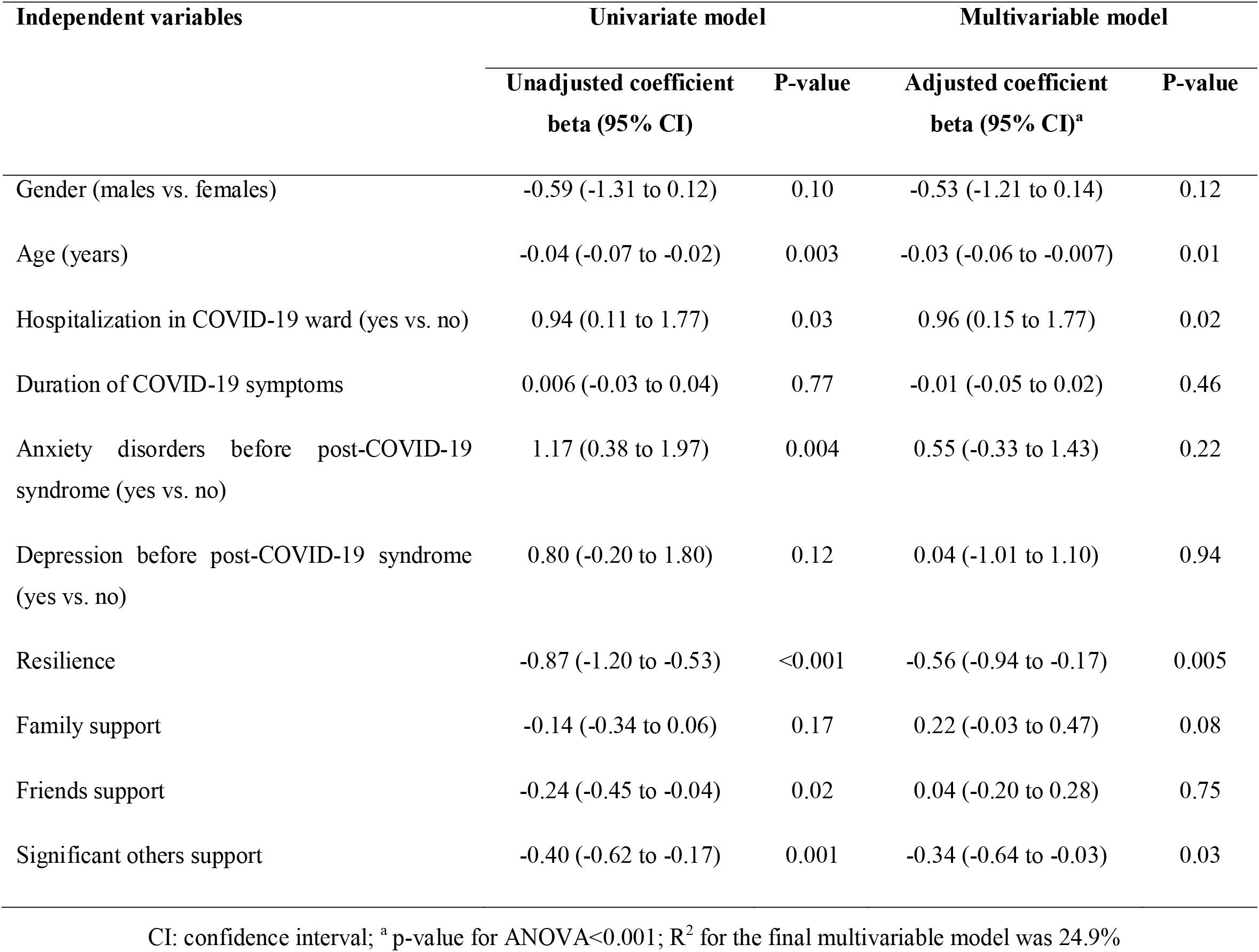
Univariate and multivariable linear regression analysis with anxiety score as the dependent variable.

| Independent variables | Univariate model |  | Multivariable model |  |
| --- | --- | --- | --- | --- |
|  | Unadjusted coefficient<br>beta (95% CI) | P-value | Adjusted coefficient<br>beta (95% CI)<sup>a</sup> | P-value |
| Gender (males vs. females) | -0.59 (-1.31 to 0.12) | 0.10 | -0.53 (-1.21 to 0.14) | 0.12 |
| Age (years) | -0.04 (-0.07 to -0.02) | 0.003 | -0.03 (-0.06 to -0.007) | 0.01 |
| Hospitalization in COVID-19 ward (yes vs. no) | 0.94 (0.11 to 1.77) | 0.03 | 0.96 (0.15 to 1.77) | 0.02 |
| Duration of COVID-19 symptoms | 0.006 (-0.03 to 0.04) | 0.77 | -0.01 (-0.05 to 0.02) | 0.46 |
| Anxiety disorders before post-COVID-19 syndrome (yes vs. no) | 1.17 (0.38 to 1.97) | 0.004 | 0.55 (-0.33 to 1.43) | 0.22 |
| Depression before post-COVID-19 syndrome (yes vs. no) | 0.80 (-0.20 to 1.80) | 0.12 | 0.04 (-1.01 to 1.10) | 0.94 |
| Resilience | -0.87 (-1.20 to -0.53) | <0.001 | -0.56 (-0.94 to -0.17) | 0.005 |
| Family support | -0.14 (-0.34 to 0.06) | 0.17 | 0.22 (-0.03 to 0.47) | 0.08 |
| Friends support | -0.24 (-0.45 to -0.04) | 0.02 | 0.04 (-0.20 to 0.28) | 0.75 |
| Significant others support | -0.40 (-0.62 to -0.17) | 0.001 | -0.34 (-0.64 to -0.03) | 0.03 |

CI: confidence interval; <sup>a</sup> p-value for ANOVA<0.001; R<sup>2</sup> for the final multivariable model was 24.9%

**Table 4.** Univariate and multivariable linear regression analysis with depression score as the dependent variable.

| Independent variables | Univariate model |  | Multivariable model |  |
| --- | --- | --- | --- | --- |
|  | Unadjusted coefficient<br>beta (95% CI) | P-value | Adjusted coefficient<br>beta (95% CI)<sup>a</sup> | P-value |
| Gender (males vs. females) | -0.81 (-1.48 to -0.13) | 0.02 | -0.78 (-1.44 to -0.12) | 0.02 |
| Age (years) | -0.03 (-0.06 to -0.004) | 0.03 | -0.02 (-0.04 to 0.008) | 0.19 |
| Hospitalization in COVID-19 ward (yes vs. no) | 0.71 (-0.09 to 1.51) | 0.08 | 0.66 (-0.14 to 1.45) | 0.10 |
| Duration of COVID-19 symptoms | 0.01 (-0.02 to 0.05) | 0.50 | -0.005 (-0.04 to 0.03) | 0.75 |
| Anxiety disorders before post-COVID-19 syndrome (yes vs. no) | 0.65 (-0.12 to 1.42) | 0.09 | 0.14 (-0.73 to 1.00) | 0.76 |
| Depression before post-COVID-19 syndrome (yes vs. no) | 0.34 (-0.62 to 1.29) | 0.48 | -0.25 (-1.29 to 0.79) | 0.64 |
| Resilience | -0.73 (-1.05 to -0.40) | <0.001 | -0.42 (-0.80 to -0.05) | 0.03 |
| Family support | -0.21 (-0.39 to -0.02) | 0.03 | 0.15 (-0.10 to 0.39) | 0.23 |
| Friends support | -0.29 (-0.48 to -0.10) | 0.003 | -0.02 (-0.26 to 0.22) | 0.89 |
| Significant others support | -0.45 (-0.66 to -0.24) | <0.001 | -0.39 (-0.69 to -0.09) | 0.01 |

CI: confidence interval; <sup>a</sup> p-value for ANOVA<0.001; R<sup>2</sup> for the final multivariable model was 19.9%

Multivariable linear regression analysis with quality of life as the dependent variable is shown in Tables 5 and 6. We found that increased resilience (adjusted beta = 0.10, 95% CI = 0.02 to 0.18) and significant others support (adjusted beta = 0.10, 95% CI = 0.04 to -0.16) was associated with increased EQ-5D-3L index value. Moreover, significant others support (adjusted beta = 5.55, 95% CI = 1.59 to 9.51) improved EQ-5D-3L VAS.

**Table 5.** Univariate and multivariable linear regression analysis with EQ-5D-3L index value as the dependent variable.

| Independent variables | Univariate model |  | Multivariable model |  |
| --- | --- | --- | --- | --- |
|  | Unadjusted coefficient<br>beta (95% CI) | P-value | Adjusted coefficient<br>beta (95% CI)<sup>a</sup> | P-value |
| Gender (males vs. females) | 0.19 (0.05 to 0.34) | 0.01 | 0.12 (-0.02 to 0.26) | 0.11 |
| Age (years) | 0.003 (-0.002 to 0.009) | 0.24 | 0.0004 (-0.005 to 0.006) | 0.88 |
| Hospitalization in COVID-19 ward (yes vs. no) | -0.06 (-0.23 to 0.11) | 0.50 | 0.008 (-0.16 to 0.18) | 0.93 |
| Duration of COVID-19 symptoms | -0.009 (-0.02 to -0.002) | 0.01 | -0.007 (-0.01 to 0.00003) | 0.05 |
| Anxiety disorders before post-COVID-19 syndrome (yes vs. no) | -0.25 (-0.42 to -0.09) | 0.002 | -0.08 (-0.26 to 0.11) | 0.40 |
| Depression before post-COVID-19 syndrome (yes vs. no) | -0.31 (-0.51 to -0.12) | 0.002 | -0.16 (-0.38 to 0.06) | 0.15 |
| Resilience | 0.15 (0.07 to 0.22) | <0.001 | 0.10 (0.02 to 0.18) | 0.01 |
| Family support | 0.03 (-0.01 to 0.07) | 0.02 | -0.04 (-0.09 to 0.01) | 0.13 |
| Friends support | 0.03 (-0.01 to 0.07) | 0.18 | -0.03 (-0.08 to 0.02) | 0.22 |
| Significant others support | 0.09 (0.04 to 0.13) | <0.001 | 0.10 (0.04 to 0.16) | 0.002 |

CI: confidence interval; <sup>a</sup> p-value for ANOVA<0.001; R<sup>2</sup> for the final multivariable model was 23.7%

**Table 6.** Univariate and multivariable linear regression analysis with EQ-5D-3L VAS as the dependent variable.

| Independent variables | Univariate model |  | Multivariable model |  |
| --- | --- | --- | --- | --- |
|  | Unadjusted coefficient<br>beta (95% CI) | P-value | Adjusted coefficient<br>beta (95% CI)<sup>a</sup> | P-value |
| Gender (males vs. females) | 9.96 (1.35 to 18.57) | 0.02 | 5.54 (-3.17 to 14.25) | 0.21 |
| Age (years) | 0.15 (-0.19 to 0.49) | 0.37 | 0.0001 (-0.34 to 0.34) | 0.99 |
| Hospitalization in COVID-19 ward (yes vs. no) | 0.63 (-9.72 to 10.98) | 0.90 | 4.53 (-5.93 to 14.99) | 0.39 |
| Duration of COVID-19 symptoms | -0.45 (-0.89 to 0.0003) | 0.05 | -0.36 (-0.80 to 0.09) | 0.11 |
| Anxiety disorders before post-COVID-19 syndrome (yes vs. no) | 14.27 (-23.92 to -4.61) | 0.004 | -6.52 (-17.92 to 4.89) | 0.26 |
| Depression before post-COVID-19 syndrome (yes vs. no) | -16.73 (-28.61 to -4.85) | 0.006 | -7.54 (-21.21 to 6.13) | 0.28 |
| Resilience | 6.35 (1.99 to 10.71) | 0.005 | 3.76 (-1.21 to 8.74) | 0.14 |
| Family support | 1.79 (-0.62 to 4.21) | 0.14 | -1.55 (-4.80 to 1.70) | 0.35 |
| Friends support | 1.38 (-1.11 to 3.88) | 0.27 | -1.82 (-4.97 to 1.34) | 0.26 |
| Significant others support | 4.63 (1.93 to 7.34) | 0.001 | 5.55 (1.59 to 9.51) | 0.006 |

CI: confidence interval; <sup>a</sup> p-value for ANOVA<0.001; R<sup>2</sup> for the final multivariable model was 15.1%

## Discussion

Literature on factors that influence mental health and quality of life in patients with post-COVID-19 syndrome is poor. The majority of studies focused only on the measurement of mental health and quality of life among patients. So far a small number of demographic and clinical characteristics of patients have been studied. To the best of our knowledge, our study is the first that investigate the role of psychosocial factors on mental health and quality of life among patients with post-COVID-19 syndrome. In particular, we explored the relationship between resilience and social support and mental health and quality of life in patients with post-COVID-19 syndrome.

Quality of life among patients with post-COVID-19 syndrome in our study was very poor. In our study, EQ-5D-3L index value was 0.36, while EQ-5D-3L index norm values based on the Greek value test is 0.916 in the 45-54 group, 0.739 in the ≥75 group, and 0.913 in all ages (Janssen et al., 2019). Moreover, EQ-5D-3L VAS in our patients was 54.1, while EQ-5D-3L VAS norm values is 78 in the 45-54 group, 54 in the ≥75 group, and 79 in all ages (Janssen et al., 2019). We compared our results with the 45-54 group since mean age of our sample 44.8 years. Thus, quality of life of our patients was even worse than that of the Greek elderly over 75 years old. Additionally, similar studies in Spain, France, USA, and China found that EQ-5D-3L VAS among patients with post-COVID-19 syndrome ranged from 64 to 80, while EQ-5D-3L index value ranged from 0.71 to 0.86 (Garrigues et al., 2020; Huang et al., 2021; Tabacof et al., 2022; Taboada et al., 2021).

Moreover, our patients experienced high levels of anxiety and depression. In particular, the proportion of our patients reporting possible major anxiety (60.7%) and depression disorder (69.7%) was much higher than reported in a meta-analysis (overall prevalence of anxiety was 23% and depression was 17%) (Premraj et al., 2022). Additionally, another meta-analysis including only studies with low/moderate risk of bias found that the overall prevalence of depression among patients recruited in the community setting was 17.3%, while the prevalence of disease among patients recruited in the hospital setting was 23.3% (European Centre for Disease Prevention and Control, 2022). Moreover, a systematic review found that the frequency of depressive symptoms ranged from 11 to 28%, while the frequency of clinically-significant depression ranged from 3 to 12% (Renaud-Charest et al., 2021).

Our results showed a significant negative relationship between resilience and anxiety and depression. In other words, the higher a patient’s resilience, the lower the anxiety and depression. Moreover, our multivariable model identified that resilience increased quality of life in our sample. Literature confirms the positive effect of resilience on individuals’ life. In particular, a meta-analysis found a significant negative correlation between resilience and psychological distress in COVID-19 patients (Jeamjitvibool et al., 2022). Similarly, a meta-analysis including patients with a somatic illness or health problem found a negative relationship between resilience and anxiety and depression (Färber & Rosendahl, 2018). Additionally, a meta-analysis including samples from the general population found a positive correlation between trait resilience and positive indicators of mental health, and a negative correlation between trait resilience and negative indicators of mental health (Hu et al., 2015). Moreover, it is well known that during the COVID-19 pandemic, resilience helped people and especially vulnerable groups (e.g., elderly or patients) to maintain good quality of life (Aldhahi et al., 2021; Javellana et al., 2022; Koivunen et al., 2022; Lipskaya-Velikovsky, 2021; Setiawan et al., 2022).

Resilience is defined as individuals’ ability to withstand or recover quickly from difficult conditions (Hu et al., 2015). In other words, resilience can be described as a defense mechanism, which gives people the ability to cope successfully with stressful experiences and bounce back from negative experiences (Lazarus, 1993). Additionally, resilience implies the flexible use of emotional resources, such as flexibility, perseverance, balance, and self-reliance for adapting to adversity (Färber & Rosendahl, 2018; Waugh et al., 2008). Therefore, resilience is crucial in promoting individuals’ positive mental health especially during the COVID-19 pandemic where mental health is compromised in several ways (e.g., lockdowns, quarantine measures, social isolation, loneliness, etc.) (Chen et al., 2022; Hessami et al., 2022; Vindegaard & Benros, 2020; Wu et al., 2021). Moreover, several studies showed that higher levels of resilience were associated with better mental health, not only in the general population but also in COVID-19 patients (MoretLTatay & Murphy, 2022; Song et al., 2021; To et al., 2022; Zhang et al., 2020, 2022). Thus, higher levels of resilience among patients with post-COVID-19 syndrome can reduce negative consequences, protect patients against adverse events, and promote patients’ ability to cope with this condition. Patients with high resilience can have a stronger capacity for self-reflection and a better tolerance of negative feelings in order to achieve a better coping with anxiety, depression, and psychological distress (Min et al., 2013).

We found that social support reduced anxiety and depression among patients with post-COVID-19 syndrome. Also, our results showed a significant positive relationship between social support and quality of life. A meta-analysis has already shown that post-COVID-19 syndrome reduces patients’ quality of life and maintains symptoms such as sleep disturbances, anxiety, depression, fatigue, and dyspnea (Malik et al., 2022). Cohort studies suggest that negative consequences of post-COVID-19 syndrome on patients’ quality of life and mental health remains even a year after the infection (Kim et al., 2022; Tabacof et al., 2022). Several studies including COVID-19 patients found that high social support had a negative association with anxiety and depression symptoms (Dai et al., 2022; Hosseini Largani et al., 2022; Kandeğer et al., 2021). Moreover, social support positively affected COVID-19 patients’ quality of life during the COVID-19 pandemic (Kesuma et al., 2022; Moodi et al., 2022). Additionally, a systematic review confirms the positive relationship between social support and people’s quality of life especially during the first wave of the pandemic (Filindassi et al., 2022).

Social support, or otherwise social network, characterizes the functioning of individuals among other people, e.g. family members, friends, significant others, and neighbors (Skalski et al., 2021). Following disasters, social support plays an essential protective role in maintaining mental health (Saltzman et al., 2020). Similarly, several studies during the pandemic suggest that people can benefit from real-life and online social support (Raude et al., 2020; Skalski et al., 2021; Wright et al., 2021; Yu et al., 2020). Social support increases belongingness and community attachment during the pandemic and that results on reduced anxiety and depressive symptoms (Alvis et al., 2022). Additionally, peer and community support groups reduces psychological distress in the era of pandemic (Nobles et al., 2020). Moreover, social support positively affects individuals’ self-efficacy during the pandemic improving their ability to cope successfully with stressful experiences, such as compliance with quarantine and isolation measures (Heffner et al., 2021; Lv et al., 2020; Tull et al., 2020). Therefore, social support is an essential psychological resource that can improve mental health and quality of life in patients with post-COVID-19 syndrome allowing them to deal successfully with the long-term psychological consequences of the disease.

Our study had several limitations. First, we used a convenience sample which is not representative of the population of patients with post-COVID-19 syndrome in Greece. For example, most of our patients were females, while only few have been hospitalized in COVID-19 intensive care unit. Moreover, our sample was obtained from the Long COVID Greece patients’ society. Thus, patients who do not belong to this society did not have the chance to participate in our study. Although we achieved the required sample size further studies with random and bigger samples can add valuable information. Second, we used self-reported tools to measure resilience, social support, anxiety, depression, and quality of life in our patients. Thus, information bias is probable in our study. Especially for anxiety and depression, we need to use valid diagnostic criteria in order get more robust results. In that case, longitudinal studies can be very helpful. Third, we assessed the independent effect of resilience and social support eliminating some confounders but other factors could be also possible confounders, e.g. socioeconomic status, educational level, family status, etc. Fourth, we performed a cross-sectional study measuring the variables at a specific time. Thus, a causal effect relationship between resilience and social support, and anxiety, depression, and quality of life cannot be established. Follow-up studies measuring changes over time could provide more valid results. Finally, we measured the impact of two psychological factors (i.e., resilience and social support) on patients’ life. Several other psychological variables (e.g., self-efficacy, mindfulness, optimism, loneliness, social integration, etc.) could affect patients’ mental health and quality of life. Thus, further studies should be performed in order to expand our knowledge on this field.

## Conclusions

Our study identified a worrying proportion of patients with post-COVID-19 syndrome with anxiety and depression symptoms. Moreover, quality of life among these patients was very poor. However, resilience and social support can be protective by reducing anxiety and depression, and improving quality of life among patients with post-COVID-19 syndrome. Prevalence of psychological problems among patients with post-COVID-19 syndrome seems to be high. Since we have to deal with a new condition our knowledge is very limited. Thus, identification of factors that influence patients’ life is crucial to reduce negative outcomes and improve quality of life.

Psychological resources, such as resilience and social support, will be important for promoting positive adaptation in case of post-COVID-19 syndrome and reducing negative symptomology. There is a need to provide on time and update psychological care services for patients with post-COVID-19 syndrome. Moreover, we should follow-up these patients for a longer period since post-COVID-19 syndrome can last for more than a year.

Patients with post-COVID-19 syndrome need time to adapt to their condition both physically and mentally. Since it is a new and unknown condition patients should develop resilience over time in order to deal effectively with the post-COVID-19 syndrome. Also, policy makers should develop and implement healthcare management programs to provide psychological support to patients.

Healthcare workers, especially clinical psychiatrists and psychologists, should be aware of the psychological needs of patients with post-COVID-19 syndrome in order to improve their mental health and quality of life. Moreover, healthcare professionals should carry out regularly follow-up observations to assess the long-term effects of post-COVID-19 syndrome.

## Data Availability

All data produced in the present study are available upon reasonable request to the authors

## Acknowledgments

The authors would like to thank the clinicians and Long COVID Greece society members who contributed their time, thoughts, and experiences to this study.

## References

Aldhahi, M. I., Akil, S., Zaidi, U., Mortada, E., Awad, S., & Al Awaji, N. (2021). Effect of Resilience on Health-Related Quality of Life during the COVID-19 Pandemic: A Cross-Sectional Study. International Journal of Environmental Research and Public Health, 18(21), 11394. https://doi.org/10.3390/ijerph182111394

Alkodaymi, M. S., Omrani, O. A., Fawzy, N. A., Shaar, B. A., Almamlouk, R., Riaz, M., Obeidat, M., Obeidat, Y., Gerberi, D., Taha, R. M., Kashour, Z., Kashour, T., Berbari, E. F., Alkattan, K., & Tleyjeh, I. M. (2022). Prevalence of post-acute COVID-19 syndrome symptoms at different follow-up periods: A systematic review and meta-analysis. Clinical Microbiology and Infection: The Official Publication of the European Society of Clinical Microbiology and Infectious Diseases, 28(5), 657–666. https://doi.org/10.1016/j.cmi.2022.01.014

Alvis, L. M., Douglas, R. D., Shook, N. J., & Oosterhoff, B. (2022). Associations between adolescents’ prosocial experiences and mental health during the COVID-19 pandemic. Current Psychology. https://doi.org/10.1007/s12144-021-02670-y

Ceban, F., Ling, S., Lui, L. M. W., Lee, Y., Gill, H., Teopiz, K. M., Rodrigues, N. B., Subramaniapillai, M., Di Vincenzo, J. D., Cao, B., Lin, K., Mansur, R. B., Ho, R. C., Rosenblat, J. D., Miskowiak, K. W., Vinberg, M., Maletic, V., & McIntyre, R. S. (2022). Fatigue and cognitive impairment in Post-COVID-19 Syndrome: A systematic review and meta-analysis. Brain, Behavior, and Immunity, 101, 93–135. https://doi.org/10.1016/j.bbi.2021.12.020

Chen, J., Zhang, S. X., Yin, A., & Yáñez, J. A. (2022). Mental health symptoms during the COVID-19 pandemic in developing countries: A systematic review and meta-analysis. Journal of Global Health, 12, 05011. https://doi.org/10.7189/jogh.12.05011

Dai, Z., Xiao, W., Wang, H., Wu, Y., Huang, Y., Si, M., Fu, J., Chen, X., Jia, M., Leng, Z., Cui, D., Dong, L., Mak, W. W. S., & Su, X. (2022). Influencing factors of anxiety and depression of discharged COVID-19 patients in Wuhan, China. PloS One, 17(11), e0276608. https://doi.org/10.1371/journal.pone.0276608

Dong, F., Liu, H., Dai, N., Yang, M., & Liu, J. (2021). A living systematic review of the psychological problems in people suffering from COVID-19. Journal of Affective Disorders, 292, 172–188. https://doi.org/10.1016/j.jad.2021.05.060

European Centre for Disease Prevention and Control. (2022). Prevalence of post COVID-19 condition symptoms: A systematic review and meta-analysis of cohort study data stratified by recruitment setting. ECDC. https://www.ecdc.europa.eu/sites/default/files/documents/Prevalence-post-COVID-19-condition-symptoms.pdf

Färber, F., & Rosendahl, J. (2018). The Association Between Resilience and Mental Health in the Somatically Ill. Deutsches Ärzteblatt International. https://doi.org/10.3238/arztebl.2018.0621

Filindassi, V., Pedrini, C., Sabadini, C., Duradoni, M., & Guazzini, A. (2022). Impact of COVID-19 First Wave on Psychological and Psychosocial Dimensions: A Systematic Review. COVID, 2(3), 273–340. https://doi.org/10.3390/covid2030022

Finstad, G. L., Giorgi, G., Lulli, L. G., Pandolfi, C., Foti, G., León-Perez, J. M., Cantero-Sánchez, F. J., & Mucci, N. (2021). Resilience, Coping Strategies and Posttraumatic Growth in the Workplace Following COVID-19: A Narrative Review on the Positive Aspects of Trauma. International Journal of Environmental Research and Public Health, 18(18), 9453. https://doi.org/10.3390/ijerph18189453

Garrigues, E., Janvier, P., Kherabi, Y., Le Bot, A., Hamon, A., Gouze, H., Doucet, L., Berkani, S., Oliosi, E., Mallart, E., Corre, F., Zarrouk, V., Moyer, J.-D., Galy, A., Honsel, V., Fantin, B., & Nguyen, Y. (2020). Post-discharge persistent symptoms and health-related quality of life after hospitalization for COVID-19. The Journal of Infection, 81(6), e4–e6. https://doi.org/10.1016/j.jinf.2020.08.029

Global Burden of Disease Long COVID Collaborators, Wulf Hanson, S., Abbafati, C., Aerts, J. G., Al-Aly, Z., Ashbaugh, C., Ballouz, T., Blyuss, O., Bobkova, P., Bonsel, G., Borzakova, S., Buonsenso, D., Butnaru, D., Carter, A., Chu, H., De Rose, C., Diab, M. M., Ekbom, E., El Tantawi, M., … Vos, T. (2022). Estimated Global Proportions of Individuals With Persistent Fatigue, Cognitive, and Respiratory Symptom Clusters Following Symptomatic COVID-19 in 2020 and 2021. JAMA, 328(16), 1604. https://doi.org/10.1001/jama.2022.18931

Heffner, J., Vives, M.-L., & FeldmanHall, O. (2021). Emotional responses to prosocial messages increase willingness to self-isolate during the COVID-19 pandemic. Personality and Individual Differences, 170, 110420. https://doi.org/10.1016/j.paid.2020.110420

Hessami, K., Romanelli, C., Chiurazzi, M., & Cozzolino, M. (2022). COVID-19 pandemic and maternal mental health: A systematic review and meta-analysis. The Journal of Maternal-Fetal & Neonatal Medicine: The Official Journal of the European Association of Perinatal Medicine, the Federation of Asia and Oceania Perinatal Societies, the International Society of Perinatal Obstetricians, 35(20), 4014–4021. https://doi.org/10.1080/14767058.2020.1843155

Hosseini Largani, M., Gorgani, F., Abbaszadeh, M., Arbabi, M., Karimpour Reyhan, S., Allameh, S. F., Shahmansouri, N., & Parsa, S. (2022). Depression, Anxiety, Perceived Stress and Family Support in COVID-19 Patients. Iranian Journal of Psychiatry, 17(3), 257–264. https://doi.org/10.18502/ijps.v17i3.9725

Hu, T., Zhang, D., & Wang, J. (2015). A meta-analysis of the trait resilience and mental health. Personality and Individual Differences, 76, 18–27. https://doi.org/10.1016/j.paid.2014.11.039

Huang, C., Huang, L., Wang, Y., Li, X., Ren, L., Gu, X., Kang, L., Guo, L., Liu, M., Zhou, X., Luo, J., Huang, Z., Tu, S., Zhao, Y., Chen, L., Xu, D., Li, Y., Li, C., Peng, L., … Cao, B. (2021). 6-month consequences of COVID-19 in patients discharged from hospital: A cohort study. Lancet (London, England), 397(10270), 220–232. https://doi.org/10.1016/S0140-6736(20)32656-8

Janssen, M. F., Szende, A., Cabases, J., Ramos-Goñi, J. M., Vilagut, G., & König, H. H. (2019). Population norms for the EQ-5D-3L: A cross-country analysis of population surveys for 20 countries. The European Journal of Health Economics, 20(2), 205–216. https://doi.org/10.1007/s10198-018-0955-5

Javellana, M., Hlubocky, F. J., Somasegar, S., Sorkin, M., Kurnit, K. C., Jani, I., Stock, E., Mills, K., Lengyel, E., & Lee, N. K. (2022). Resilience in the Face of Pandemic: The Impact of COVID-19 on the Psychologic Morbidity and Health-Related Quality of Life Among Women With Ovarian Cancer. JCO Oncology Practice, 18(6), e948–e957. https://doi.org/10.1200/OP.21.00514

Jeamjitvibool, T., Duangchan, C., Mousa, A., & Mahikul, W. (2022). The Association between Resilience and Psychological Distress during the COVID-19 Pandemic: A Systematic Review and Meta-Analysis. International Journal of Environmental Research and Public Health, 19(22), 14854. https://doi.org/10.3390/ijerph192214854

Kandeğer, A., Aydın, M., AltinbaŞ, K., Cansız, A., Tan, Ö., Tomar Bozkurt, H., Eğilmez, Ü., Tekdemir, R., Şen, B., Aktuğ Demir, N., Sümer, Ş., Ural, O., Yormaz, B., Ergün, D., Tülek, B., & Kanat, F. (2021). Evaluation of the relationship between perceived social support, coping strategies, anxiety, and depression symptoms among hospitalized COVID-19 patients. International Journal of Psychiatry in Medicine, 56(4), 240–254. https://doi.org/10.1177/0091217420982085

Karekla, M., Pilipenko, N., & Feldman, J. (2012). Patient Health Questionnaire: Greek language validation and subscale factor structure. Comprehensive Psychiatry, 53(8), 1217–1226. https://doi.org/10.1016/j.comppsych.2012.05.008

Kesuma, E., Purwadi, H., Sanjaya Putra, D., & Pranata, S. (2022). Social support Improved the quality of life among Covid-19 Survivors in Sumbawa. International Journal of Nursing and Health Services, 4(5), 319–327.

Kim, Y., Bitna-Ha, Kim, S.-W., Chang, H.-H., Kwon, K. T., Bae, S., & Hwang, S. (2022). Post-acute COVID-19 syndrome in patients after 12 months from COVID-19 infection in Korea. BMC Infectious Diseases, 22(1), 93. https://doi.org/10.1186/s12879-022-07062-6

Koivunen, K., Portegijs, E., Sillanpää, E., Eronen, J., Kokko, K., & Rantanen, T. (2022). Maintenance of high quality of life as an indicator of resilience during COVID-19 social distancing among community-dwelling older adults in Finland. Quality of Life Research: An International Journal of Quality of Life Aspects of Treatment, Care and Rehabilitation, 31(3), 713–722. https://doi.org/10.1007/s11136-021-03002-0

Kroenke, K., Spitzer, R. L., Williams, J. B. W., & Löwe, B. (2009). An ultra-brief screening scale for anxiety and depression: The PHQ-4. Psychosomatics, 50(6), 613–621. https://doi.org/10.1176/appi.psy.50.6.613

Kyriazos, T. A., Stalikas, A., Prassa, K., Galanakis, M., Yotsidi, V., & Lakioti, A. (2018). Psychometric Evidence of the Brief Resilience Scale (BRS) and Modeling Distinctiveness of Resilience from Depression and Stress. Psychology, 09(07), 1828–1857. https://doi.org/10.4236/psych.2018.97107

Labrague, L. J. (2021). Psychological resilience, coping behaviours and social support among health care workers during the COVID-19 pandemic: A systematic review of quantitative studies. Journal of Nursing Management, 29(7), 1893–1905. https://doi.org/10.1111/jonm.13336

Lazarus, R. S. (1993). From psychological stress to the emotions: A history of changing outlooks. Annual Review of Psychology, 44, 1–21. https://doi.org/10.1146/annurev.ps.44.020193.000245

Lipskaya-Velikovsky, L. (2021). COVID-19 Isolation in Healthy Population in Israel: Challenges in Daily Life, Mental Health, Resilience, and Quality of Life. International Journal of Environmental Research and Public Health, 18(3), 999. https://doi.org/10.3390/ijerph18030999

Long COVID Europe. (2023, February 4). https://longcovideurope.org/

Long COVID Greece. (2023, February 4). https://longcovidgreece.gr/

Lorah, J. (2018). Effect size measures for multilevel models: Definition, interpretation, and TIMSS example. Large-Scale Assessments in Education, 6(1), 8. https://doi.org/10.1186/s40536-018-0061-2

Lv, Y., Yao, H., Xi, Y., Zhang, Z., Zhang, Y., Chen, J., Li, J., Li, J., Wang, X., & Luo, G. H. (2020). Social Support Protects Chinese Medical Staff from Suffering Psychological Symptoms in COVID-19 Defense. SSRN Electronic Journal. https://doi.org/10.2139/ssrn.3559617

Malik, P., Patel, K., Pinto, C., Jaiswal, R., Tirupathi, R., Pillai, S., & Patel, U. (2022). PostLacute COVIDL19 syndrome (PCS) and healthLrelated quality of life (HRQoL)—A systematic review and metaLanalysis. Journal of Medical Virology, 94(1), 253–262. https://doi.org/10.1002/jmv.27309

Min, J.-A., Yoon, S., Lee, C.-U., Chae, J.-H., Lee, C., Song, K.-Y., & Kim, T.-S. (2013). Psychological resilience contributes to low emotional distress in cancer patients. Supportive Care in Cancer: Official Journal of the Multinational Association of Supportive Care in Cancer, 21(9), 2469–2476. https://doi.org/10.1007/s00520-013-1807-6

Moodi, M., Sharifzadeh, G., & Baghernejad, F. (2022). Evaluation of Perceived Social Support Status and Quality of Life in Improved COVID-19 Patients in Birjand, Iran. Modern Care Journal, 19(1). https://doi.org/10.5812/modernc.120955

Moret□Tatay, C., & Murphy, M. (2022). Anxiety, resilience and local conditions: A CROSS□CULTURAL investigation in the time of Covid□19. International Journal of Psychology, 57(1), 161–170. https://doi.org/10.1002/ijop.12822

Nasserie, T., Hittle, M., & Goodman, S. N. (2021). Assessment of the Frequency and Variety of Persistent Symptoms Among Patients With COVID-19: A Systematic Review. JAMA Network Open, 4(5), e2111417. https://doi.org/10.1001/jamanetworkopen.2021.11417

Nobles, J., Martin, F., Dawson, S., Moran, P., & Savovic, J. (2020). The Potential Impact of COVID-19 on Mental Health Outcomes and the Implications for Service Solutions. National Institute for Health Research, University of Bristol.

Premraj, L., Kannapadi, N. V., Briggs, J., Seal, S. M., Battaglini, D., Fanning, J., Suen, J., Robba, C., Fraser, J., & Cho, S.-M. (2022). Mid and long-term neurological and neuropsychiatric manifestations of post-COVID-19 syndrome: A meta-analysis. Journal of the Neurological Sciences, 434, 120162. https://doi.org/10.1016/j.jns.2022.120162

Rabin, R., & de Charro, F. (2001). EQ-5D: A measure of health status from the EuroQol Group. Annals of Medicine, 33(5), 337–343. https://doi.org/10.3109/07853890109002087

Raude, J., Lecrique, J.-M., Lasbeur, L., Leon, C., Guignard, R., du Roscoät, E., & Arwidson, P. (2020). Determinants of Preventive Behaviors in Response to the COVID-19 Pandemic in France: Comparing the Sociocultural, Psychosocial, and Social Cognitive Explanations. Frontiers in Psychology, 11, 584500. https://doi.org/10.3389/fpsyg.2020.584500

Renaud-Charest, O., Lui, L. M. W., Eskander, S., Ceban, F., Ho, R., Di Vincenzo, J. D., Rosenblat, J. D., Lee, Y., Subramaniapillai, M., & McIntyre, R. S. (2021). Onset and frequency of depression in post-COVID-19 syndrome: A systematic review. Journal of Psychiatric Research, 144, 129–137. https://doi.org/10.1016/j.jpsychires.2021.09.054

Saltzman, L. Y., Hansel, T. C., & Bordnick, P. S. (2020). Loneliness, isolation, and social support factors in post-COVID-19 mental health. Psychological Trauma: Theory, Research, Practice, and Policy, 12(S1), S55–S57. https://doi.org/10.1037/tra0000703

Setiawan, T., Wardani, R., & Theresia, E. (2022). The conditional effect of family resilience on family quality of life during the Covid-19 pandemic. F1000Research, 11, 1279. https://doi.org/10.12688/f1000research.125852.3

Shah, W., Hillman, T., Playford, E. D., & Hishmeh, L. (2021). Managing the long term effects of covid-19: Summary of NICE, SIGN, and RCGP rapid guideline. BMJ, n136. https://doi.org/10.1136/bmj.n136

Shanbehzadeh, S., Tavahomi, M., Zanjari, N., Ebrahimi-Takamjani, I., & Amiri-arimi, S. (2021). Physical and mental health complications post-COVID-19: Scoping review. Journal of Psychosomatic Research, 147, 110525. https://doi.org/10.1016/j.jpsychores.2021.110525

Skalski, S., Uram, P., Dobrakowski, P., & Kwiatkowska, A. (2021). The link between ego-resiliency, social support, SARS-CoV-2 anxiety and trauma effects. Polish adaptation of the Coronavirus Anxiety Scale. Personality and Individual Differences, 171, 110540. https://doi.org/10.1016/j.paid.2020.110540

Smith, B. W., Dalen, J., Wiggins, K., Tooley, E., Christopher, P., & Bernard, J. (2008). The brief resilience scale: Assessing the ability to bounce back. International Journal of Behavioral Medicine, 15(3), 194–200. https://doi.org/10.1080/10705500802222972

Song, S., Yang, X., Yang, H., Zhou, P., Ma, H., Teng, C., Chen, H., Ou, H., Li, J., Mathews, C. A., Nutley, S., Liu, N., Zhang, X., & Zhang, N. (2021). Psychological Resilience as a Protective Factor for Depression and Anxiety Among the Public During the Outbreak of COVID-19. Frontiers in Psychology, 11, 618509. https://doi.org/10.3389/fpsyg.2020.618509

Tabacof, L., Tosto-Mancuso, J., Wood, J., Cortes, M., Kontorovich, A., McCarthy, D., Rizk, D., Rozanski, G., Breyman, E., Nasr, L., Kellner, C., Herrera, J. E., & Putrino, D. (2022). Post-acute COVID-19 Syndrome Negatively Impacts Physical Function, Cognitive Function, Health-Related Quality of Life, and Participation. American Journal of Physical Medicine & Rehabilitation, 101(1), 48–52. https://doi.org/10.1097/PHM.0000000000001910

Taboada, M., Moreno, E., Cariñena, A., Rey, T., Pita-Romero, R., Leal, S., Sanduende, Y., Rodríguez, A., Nieto, C., Vilas, E., Ochoa, M., Cid, M., & Seoane-Pillado, T. (2021). Quality of life, functional status, and persistent symptoms after intensive care of COVID-19 patients. British Journal of Anaesthesia, 126(3), e110–e113. https://doi.org/10.1016/j.bja.2020.12.007

Theofilou, P. (2015). Translation and cultural adaptation of the Multidimensional Scale of Perceived Social Support for Greece. Health Psychology Research, 3(1). https://doi.org/10.4081/hpr.2015.1061

To, Q. G., Vandelanotte, C., Cope, K., Khalesi, S., Williams, S. L., Alley, S. J., Thwaite, T. L., Fenning, A. S., & Stanton, R. (2022). The association of resilience with depression, anxiety, stress and physical activity during the COVID-19 pandemic. BMC Public Health, 22(1), 491. https://doi.org/10.1186/s12889-022-12911-9

Tull, M. T., Edmonds, K. A., Scamaldo, K. M., Richmond, J. R., Rose, J. P., & Gratz, K. L. (2020). Psychological Outcomes Associated with Stay-at-Home Orders and the Perceived Impact of COVID-19 on Daily Life. Psychiatry Research, 289, 113098. https://doi.org/10.1016/j.psychres.2020.113098

Vindegaard, N., & Benros, M. E. (2020). COVID-19 pandemic and mental health consequences: Systematic review of the current evidence. Brain, Behavior, and Immunity, 89, 531–542. https://doi.org/10.1016/j.bbi.2020.05.048

Waugh, C. E., Fredrickson, B. L., & Taylor, S. F. (2008). Adapting to life’s slings and arrows: Individual differences in resilience when recovering from an anticipated threat. Journal of Research in Personality, 42(4), 1031–1046. https://doi.org/10.1016/j.jrp.2008.02.005

World Health Organization. (2023a). A clinical case definition of post COVID-19 condition by a Delphi consensus, 6 October 2021. https://www.who.int/publications/i/item/WHO-2019-nCoV-Post_COVID-19_condition-Clinical_case_definition-2021.1

World Health Organization. (2023b). At least 17 million people in the WHO European Region experienced long COVID in the first two years of the pandemic; millions may have to live with it for years to come. https://www.who.int/europe/news/item/13-09-2022-at-least-17-million-people-in-the-who-european-region-experienced-long-covid-in-the-first-two-years-of-the-pandemic--millions-may-have-to-live-with-it-for-years-to-come

Wright, L., Steptoe, A., & Fancourt, D. (2021). Are adversities and worries during the COVID-19 pandemic related to sleep quality? Longitudinal analyses of 46,000 UK adults. PloS One, 16(3), e0248919. https://doi.org/10.1371/journal.pone.0248919

Wu, T., Jia, X., Shi, H., Niu, J., Yin, X., Xie, J., & Wang, X. (2021). Prevalence of mental health problems during the COVID-19 pandemic: A systematic review and meta-analysis. Journal of Affective Disorders, 281, 91–98. https://doi.org/10.1016/j.jad.2020.11.117

Yfantopoulos, J. (1999). Quality of life measurement and health production in Greece. In EuroQol plenary meeting. Discussion papers (pp. 100–114). Uni-Verlag Witte.

Yu, H., Li, M., Li, Z., Xiang, W., Yuan, Y., Liu, Y., Li, Z., & Xiong, Z. (2020). Coping style, social support and psychological distress in the general Chinese population in the early stages of the COVID-19 epidemic. BMC Psychiatry, 20(1), 426. https://doi.org/10.1186/s12888-020-02826-3

Zhang, J., Wang, Y., Zhou, M., & Ke, J. (2022). Community resilience and anxiety among Chinese older adults during COVID L19: The moderating role of trust in local government. Journal of Community & Applied Social Psychology, 32(3), 411–422. https://doi.org/10.1002/casp.2563

Zhang, J., Yang, Z., Wang, X., Li, J., Dong, L., Wang, F., Li, Y., Wei, R., & Zhang, J. (2020). The relationship between resilience, anxiety and depression among patients with mild symptoms of COVIDL19 in China: A crossLsectional study. Journal of Clinical Nursing, 29(21–22), 4020–4029. https://doi.org/10.1111/jocn.15425

Zimet, G. D., Dahlem, N. W., Zimet, S. G., & Farley, G. K. (1988). The Multidimensional Scale of Perceived Social Support. Journal of Personality Assessment, 52(1), 30–41. https://doi.org/10.1207/s15327752jpa5201_2

